# Growing up without violence (GWV): Results of a cluster randomised trial of a school-based intervention preventing adolescent sexual abuse and exploitation in Brazil

**DOI:** 10.64898/2026.01.21.26344579

**Authors:** Ligia Kiss, Marto Leal, Ana Paula Portella, Heloína Paiva, Agnese Iuliano, Malini Pires, Helen Shipman, Elizabeth Anderson, Yuki Lo, Ligia Kerr, Carl Kendall, Dario Brito, Marina Barros, Baptiste Leurent

## Abstract

**Introduction:** Child sexual abuse and exploitation (CSAE) are pervasive problems that significantly affect the health of children and adolescents. This paper presents the results of a cluster randomised controlled trial (cRCT) evaluating a school-based intervention designed to prevent CSAE in Brazil through education.

**Methods:** We conducted a two-arm cRCT with parallel assignment in two Brazilian municipalities. Sixty schools were randomly allocated to intervention and control arms (30 per group) using stratified randomisation based on municipality and school type (municipal and state-run). We invited 50 randomly selected students, aged 12 to 17, from each school to complete surveys at baseline and endline. Teachers enrolled in GWV training completed an implementation questionnaire. Our primary analysis focused on a cross-sectional comparison of students’ CSAE risk knowledge between control and intervention schools.

**Results:** We found no statistically significant difference in adolescents’ CSAE risk knowledge between the study arms (adjusted mean difference: 0.02, 95% CI −0.19 to 0.23, p = 0.87). There was borderline evidence of interaction by gender, with a greater difference in knowledge scores among girls (p=0.032). No significant association was found between the number of GWV components implemented and knowledge scores.

**Discussion:** Several factors may contribute to this result: the intervention might not have been implemented with sufficient fidelity or intensity, the true effect of GWV could be minimal and below the study’s detection threshold, the intervention may have influenced outcomes that were not measured, or the intervention may simply lack effectiveness. Further rigorous evaluations and implementation studies are essential to ensure that resources are directed toward effective strategies.

**Trial registration:** The American Economic Association’s registry for randomised controlled trials (AEA RCT Registry). RCT ID: AEARCTR-0011299, https://www.socialscienceregistry.org/trials/11299

**Funding:** Funding for this study was granted by the United States Department of State under the terms of Cooperative Agreement No. SSJTIP20CA0026, through the Human Trafficking Research Initiative, managed by Innovations for Poverty Action

## INTRODUCTION

Child sexual abuse and exploitation (CSAE) are associated with severe negative health impacts, including early pregnancy, sexually transmitted infections (STIs), gynaecological problems, and violence-related injuries. The mental health consequences of CSAE can be profound and long-lasting, and include anxiety, depression, post-traumatic stress disorder, developmental problems, suicidal behaviour, self-harm, and substance misuse (1–3). Many of these problems can persist into adulthood. Nonetheless, despite adolescents’ high risk of sexual abuse and exploitation compared to other age groups, this public health agenda remains underdeveloped across the world (3).

The World Health Organisation (WHO) defines child sexual abuse as actual or threatened physical intrusion of a sexual nature, whether by force or under conditions of inequality or coercion. Sexual exploitation involves the abuse of a position of vulnerability, power, or trust, for sexual purposes (4). Child sexual exploitation encompasses any form of transactional sex, including sex trafficking, the sale of images or videos depicting the sexual assault of children, sex tourism, and paid sexual performances such as livestreaming (5).

The global prevalence of sexual violence against children is estimated at 18·9% (95% uncertainty interval [UI] 16·0–25·2) for females and 14·8% (9·5–23·5) for males in 2023 (6). A recent study involving a national sample of Brazilian students aged 13 to 17 reported prevalence rates of 14.6% for child sexual abuse and 6.3% for rape (7). Research in the Recife Metropolitan Region (RMR), our study site, estimates that 9.62% (5.44-18.83%) of girls under 18 were commercially sexually exploited between 2019 and 2022. Of these adolescents, 78.6% were still enrolled in school when first exploited (8, 9).

Evidence suggests that the health burden of CSAE can be prevented through targeted investments in public health approaches that emphasise education and awareness, early intervention, and adolescent-led responses (3, 10). A meta-analysis of 24 studies found that school-based education programs aimed at addressing sexual abuse can enhance children’s knowledge of CSAE, improve self-protective behaviours, increase the likelihood of disclosure, and reduce feelings of self-blame. However, there is limited evidence regarding the effectiveness of these interventions in low- and middle-income countries (11).

This paper presents findings from the Growing Up Without Violence (GWV) trial, which evaluated the effectiveness of the GWV curriculum as a school-based prevention program for child sexual abuse and exploitation (CSAE). Our study aimed to assess the intervention’s impact on students’ knowledge of the risks associated with sexual abuse and exploitation, as well as their self-protective behaviours.

### Growing up Without Violence

For more than two decades, Brazil has been promoting policies to address sexual violence and sexual exploitation of children and adolescents (12). The country is equipped with local and regional social service offices to address social risks and rights violations, including violence against children (13). Although these resources exist, schools often lack the protocols, training, and support to detect, manage, and refer cases of CSAE (14–16). Growing-up Without Violence (GWV) aims to address this gap through a school-based curriculum to reduce students’ risks of sexual abuse and exploitation and increase their reporting skills.

GWV is a large-scale intervention in Brazil aimed at preventing CSAE. The intervention was developed and is implemented by Canal Futura, a non-profit arm of the Roberto Marinho Foundation, the largest media conglomerate in Brazil. GWV was initially developed in 2009 through thematic fora and consultations with experts and adolescent CSAE survivors. These fora periodically revise strategies, informing priorities and best implementation practices, and contributing to content development. The intervention’s curriculum aims to enhance students’ understanding of CSAE risks, self-protection strategies, and reporting mechanisms, while also equipping teachers with the skills needed to identify and respond effectively to cases. GWV assumes that adolescents equipped with this knowledge will be better able to recognise and respond effectively to CSAE risks. The intervention targets students aged 7 and older enrolled in public schools in Brazil and is tailored to different age groups and their developmental stages.

To implement the intervention, Canal Futura leverages its partnerships with Brazilian local authorities, who identify the target schools that will receive the intervention. Typically, these are public schools managed by these authorities at both the municipal and state levels. In the research sites for this study, all eligible schools were invited to participate. Eligible schools included all publicly funded institutions with students aged 12 to 17 enrolled.

The relevant local authority requested that each school select two teachers to participate in capacity-building sessions. These sessions focus on themes presented in the intervention’s audiovisual content, including family-based child sexual abuse, sexual exploitation, victim identification, active listening, reporting CSAE, intersectoral victim support, children’s emotional health (including self-harm risks), and discrimination related to gender identity and sexual orientation. Supplementary Material 1 provides a summary of the intervention’s content, target population, and materials.

All GWV trainers have a professional background in education and sexual violence prevention and undergo intensive training on the intervention. This training begins with an induction meeting, followed by five two-hour capacity-building sessions in which Canal Futura enhances participants’ knowledge about CSAE and provides support for planning and implementing GWV resources in school classrooms. During the planning, implementation, and feedback stages, trainees receive ongoing support and share their experiences through a social media group managed by Canal Futura. School staff ultimately make the final decisions on GWV content and frequency, based on their priorities, capacity, and available resources. In the final training session, participants engage in a workshop to share their experiences with the curriculum and discuss long-term plans for project sustainability.

## MATERIALS AND METHODS

### Study design

The GWV Trial is a two-arm parallel repeated cross-sectional Cluster Randomised Controlled Trial (cRCT) of the GWV curriculum, a school-based complex social intervention to prevent CSAE.

### Patient and Public Involvement

The study was planned and conducted in collaboration with local authorities, school managers, and teachers, who were initially engaged in presentations and discussions in Cabo de Santo Agostinho in 2023. Many of the authors of the manuscript were present during this event. Subsequent meetings were held online. Research questions and study outcomes were co-developed with Canal Futura and the Freedom Fund, drawing on their experience with schools and staff training. School managers played a crucial role in obtaining consent from parents. The methods for the study were specified in the research call by IPA and members of the public. Members of the public were not involved in determining the plans for disseminating the study.

### Schools and participant inclusion criteria and selection

Public schools in the municipalities of Cabo de Santo Agostinho and Jaboatão dos Guareligarapes (hereafter referred to as Cabo and Jaboatão, respectively), with students aged 12 to 17, served as the units of randomization for the cRCT. Both municipalities are located in the Metropolitan Region of Recife, in the northeastern Brazilian state of Pernambuco, and are recognized as hotspots for adolescent sexual exploitation in Brazil (17). Cabo is home to the largest port in Brazil Northeast, the Suape Port. As a transit point for sexual tourism, the municipality experiences high rates of adolescent sexual exploitation and child drug use (18). Jaboatão is one of the three municipalities with the highest population density in Pernambuco and is also recognized for having a high incidence of adolescent sexual exploitation. (19).

All schools partaking in the study were publicly funded and followed the normative national educational curricula. From each selected school, two independent cross-sectional samples of students were invited to participate in the baseline and endline surveys. Eligible students were those in the final two years of primary/elementary school or in secondary school (ages 12 to 17) who could understand the survey questions and whose parents did not withdraw consent for their participation in the survey.

### Randomisation and masking

The trial statistician prepared a randomisation program using a random number generator in Stata (20). Allocation between the two trial arms was with a 1:1 ratio stratified by municipality and school type, composing three strata: i) Cabo, municipal schools, ii) Cabo, state schools, and iii) Jaboatão, state schools. The trial coordinator ran the randomisation program and identified which of the 60 schools were allocated to the GWV intervention or waitlist control.

The trial coordinator informed Canal Futura of the allocation. Schools and students were not masked to the allocation but were not informed until after the baseline survey (described below). The principal investigator, the trial statistician, and the research team were blinded to study allocation. Unmasking occurred once all the endline data were collected and processed, and the statistical analysis plan was approved and published.

Implementation data were processed after unmasking.

### Intervention and control

After being informed about the schools randomised to the GWV arm, Canal Futura approached them to schedule the training sessions.

Schools in the control arm did not receive the intervention and adhered to standard national guidelines, which typically require schools to implement activities focused on the week of May 18th, the Brazilian National Day to Combat Abuse and Sexual Exploitation of Children and Adolescents. Government guidelines mandate that Brazilian schools promote activities during this week to raise awareness about CSAE. However, these schools do not receive any training or guidance on the content or implementation of educational activities. Each intervention school selected two teachers to receive training. Participating teachers attended an induction meeting where Canal Futura’s trained experts outlined GWV’s aims, principles, and curriculum content. The selected teachers were trained on the GWV material, following Canal Futura’s standard methodology, as previously described. To ensure that children in the control arm also received the information covered by the GWV intervention, Canal Futura offered the intervention to schools in the control arm after the study’s endline.

### Data collection

Students from the participating schools were invited to complete a survey at baseline in May 2024 (just after randomisation, and before the intervention was implemented), and at endline in September (approximately 4 months after baseline).

For both surveys, students were randomly selected from lists of all eligible participants at each school. The average number of eligible students per school was 273, ranging from 27 to 690. We piloted the survey in one municipal school in Cabo and one state school in Jaboatão. Given the high levels of student absenteeism observed in the baseline pilot results (approximately 28% of students in the selected sample), we randomly selected 70 students from schools with larger enrolments (n > 70), intending to interview 50. All students from the four schools with fewer than 50 eligible students were invited.

School managers supported the research team in implementing an opt-out informed consent process with all parents of selected students. Parents were provided with information about the study and given the opportunity to withdraw their child’s participation. School managers contacted eligible parents through their usual communication channels (primarily WhatsApp groups), where they shared study information, responded to queries with support from the research team, and offered the option to opt out. Lists of parents who declined participation were subsequently shared with the research team, and the corresponding students were removed from the sampling frame.

Participating students received an electronic information sheet on the survey tablets explaining that the study included questions about potentially distressing past experiences and advising them not to participate if they believed this could cause discomfort. At both baseline and endline, fieldwork supervisors briefed students on the study procedures and ethical safeguards, emphasising voluntary participation, the right to refuse or withdraw, and the confidentiality and anonymity protections. The survey tablet was programmed to allow access to the questionnaire only after students provided active consent.

Fieldwork supervisors and researchers were trained using an adapted version of an adapted version of the WHO ethics protocols for research on violence against women (21) and on human trafficking (21), as well as guidelines for ethical research with children (22). Training covered interviewing procedures, safety and confidentiality, responding to sample, referral pathways, and safeguarding of both participants and research staff. A specialised psychologist conducted an additional session on responding to children who exhibited distress or required immediate assistance.

Students completed self-administered surveys lasting 30-45 minutes. Fieldwork researchers ensured privacy, assisted with understanding the content, and provided individual support before and after the survey (maintaining a ratio of one researcher to five participants). The research team ensured that interviews were conducted in a private and safe environment, that participants fully understood the consent procedures, and that they freely agreed to participate and provided voluntary written assent. Details of the trial’s ethical procedures can be found in the protocol paper (23).

Teachers who participated in the GWV training were also invited to fill in a survey about their implementation of the GWV curriculum. Two staff members in the trial fieldwork team implemented this survey immediately before the endline. Researchers sent the monitoring questionnaire to teachers in intervention schools through email and phone. They followed up by phone with those who did not initially respond.

### Outcomes

The primary outcome of the cRCT was children’s knowledge of inappropriate sexual advances and acts, including grooming. We utilized the revised Children’s Knowledge of Abuse Questionnaire (CKAQ-R), which has been validated in Portuguese (24), to assess children’s understanding of sexual abuse prevention concepts at both baseline and endline (25). Children responded to 12 items, indicating whether they agreed, disagreed, or were unsure if the described behaviour was abusive. We scored the responses based on evidence-based guidelines and definitions of abuse. Correct identifications of abusive behaviour received 1 point, while incorrect responses (i.e., failing to identify abuse or mislabelling appropriate behaviour as abuse) and "unsure" responses received 0 points. The total CKAQ-R score was derived by summing all responses, yielding a range of 0 to 12, with higher scores indicating greater knowledge of abuse.

Additionally, we measured the effect of GWV on three secondary outcomes: i. knowledge of risks associated with online sex; ii. intention to seek help in cases of sexual abuse or exploitation, and iii. recall of recent exposure to school-based CSAE prevention activities. Knowledge of risks associated with online sex was measured by fifteen questions adapted to themes explored in the GWV curriculum, such as sexting, cyber grooming, and online sexual exploitation. Correct answers were scored as 1 point, and incorrect and "unsure" responses as 0 points. The total knowledge score was derived by summing all questions, with higher scores indicating greater knowledge (range 0 to 15). We assessed intention to seek help in cases of sexual abuse or exploitation through positive responses to the question "Would you seek help in a bad situation where you had a nude leaked or had your intimacy exposed or were sexually harassed online?". This outcome was scored 1 if the student answered “yes” and as 0 if they answered “no” or “unsure”. Recall of recent exposure to school-based CSAE prevention activities was determined by responses to the question "Have you ever participated in teaching activities, workshops, or classes on how to prevent or react to situations in which someone wants to touch your body in a way that bothers you or that you don’t want?”. This outcome was scored as 1 if the student answered “yes” and as 0 if they answered “no” or “unsure”. Further details on the outcomes’ definitions are available in the published protocol (26) and the Statistical Analysis Plans (26).

### Other measures

Other measures captured during baseline and endline included students’ characteristics, such as age, gender, and their experience of sexual abuse and exploitation. Items on sexual exploitation were based on the items used in the Violence Against Children (VAC) and Youth Surveys (27) and research by Dunkle, Jewkes (28). These items were refined through recommendations by Wamoyi, Ranganathan (29). We measured both transactional sex and sex work. We used questions from an adapted version of the questionnaire for the Project Sao Paulo for the social development of children and adolescents (SP-PROSO) (30).

Data on intervention implementation included the number of teachers registered to attend the GWV trainings and data from the teachers’ survey, including how many ‘components’ (i.e., specific material provided by GWV or topics covered as part of the training) they had already used in class. We used the number of components to define a level of GWV implementation (dose). We categorised schools into three groups: no data (school did not implement the intervention, or no teachers completed the implementation survey); lower implementation (teachers reporting an average of components used of 3 or less); higher implementation (average above 3. We considered components implemented through the reported use of specific programme guidelines and learning materials for student activities (e.g., educational GWV kit, audiovisual series, workbooks on GWV themes, campaign materials, and case studies).

### Sample size

The study was powered to detect a small to medium standardised difference of 0.25 on the CKAQ-R at the endline. Assuming an intra-cluster correlation coefficient (ICC) of 0.07, 60 schools (30 per arm) with 50 students sampled in each school provide 90% power to detect an effect size of 0.25 at the 2-sided alpha level of 5%.

### Statistical analysis

Details of the statistical analysis methods are reported in the Statistical Analysis Plan (26). Descriptive statistics of schools and students’ characteristics at baseline are reported. The primary estimand of interest is the mean difference in CKAQ-R score at endline for schools randomised to receive the GWV curriculum compared to those that did not. We compared the primary outcome (CKAQ-R score at endline) between arms using a cluster-level summary approach, deriving the mean score for each school, then estimating the mean difference between arms using a linear regression (31, 32). We adjusted the analysis by the schools’ mean CKAQ-R score at baseline and stratified by randomisation strata. An equal weight was given to each school. We compared the continuous secondary outcome (knowledge of risks associated with online sex) similarly, adjusting for the equivalent outcome at baseline. For the binary outcomes (seeking help in case of online abuse and participation in sexual abuse prevention activity), we reported both the risk difference and risk ratio, estimated using a cluster-level approach stratified by randomisation strata. We conducted the hypothesis tests based on the risk differences. The risk ratios were based on the ratio of geometric means (32). We conducted a comparative analysis according to the randomisation arm, regardless of the intervention received (intention-to-treat).

We then estimated the effect modification between students’ age and gender on the primary outcome, using linear mixed models with an interaction term between arm and the baseline covariates (age or gender). The models included a random-effect by school and were adjusted by baseline mean CKAQ-R and randomisation strata.

Finally, we conducted a pre-specified exploratory analysis examining the association between the intervention ‘dose’ and the primary outcome in the schools randomised to the intervention. We categorised the schools into three groups based on teachers’ responses to the implementation survey (see ‘other measures’) and performed an overall test for differences using linear regression. This analysis compared the school-level data (mean endline CKAQ-R score), adjusting for strata and baseline CKAQ-R score. We conducted all analyses in Stata v18 (20) and used the clan command for cluster-level analysis (32).

## RESULTS

### Recruitment and randomisation

We identified 82 public schools with students aged 12 to 17 in Cabo and Jaboatão municipalities (Figure 1). We enrolled all 31 municipal schools from Cabo in the trial, along with 29 randomly selected state schools from both Cabo and Jaboatão. All 60 schools were randomized, with half (n=30) allocated to receive the GWV training and the other half (n=30) assigned to the control arm. Figure 1 shows the trial flow chart.

**Figure 1.**
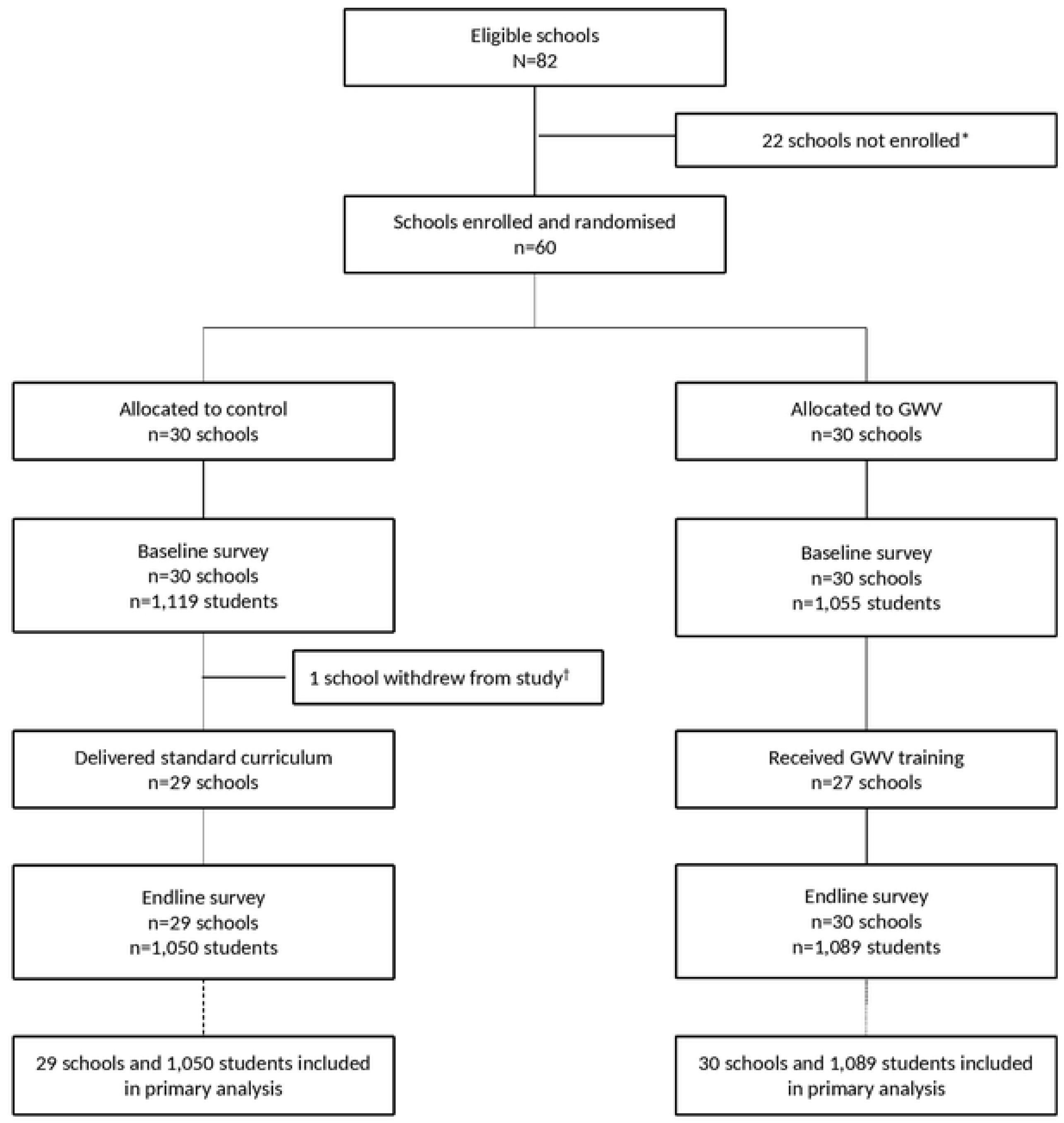
Trial flow chart. ^*^ All 31 municipal schools from Cabo were enrolled, and 29 from 51 state schools in Cabo and Jaboatão were randomly selected to reach the sample size of 60 schools. ^†^ One school withdrew from the study after baseline because it did not feel the research topic was suitable for their students. Note: school randomisation was conducted before the baseline survey for logistical constraints, but no intervention had been delivered by the time of the baseline survey. Students and school staff were unaware of the allocation.

### Baseline

We conducted the baseline survey between April and May 2024 with 2,174 students (36.2 students per school) (Table 1).

**Table 1.** Baseline characteristics.

|  | Control | GWV | All |
| --- | --- | --- | --- |
| **School characteristics** |  |  |  |
| **Number of schools** | 30 | 30 | 60 |
| **Strata** |  |  |  |
| Cabo de Santo Agostinho – Municipal schools | 14 (46.7%) | 14 (46.7%) | 28 (46.7%) |
| Cabo de Santo Agostinho – State schools | 4 (13.3%) | 4 (13.3%) | 8 (13.3%) |
| Jaboatão dos Guararapes – State schools | 12 (40.0%) | 12 (40.0%) | 24 (40.0%) |
| **Number of eligible students per school\*** | 278 (153) | 268 (177) | 273 (164) |
| **Number of respondents per school** | 37.3 (9.4) | 35.2 (9.8) | 36.2 (9.6) |
| **Student characteristics** |  |  |  |
| **Number of student respondents** | 1,119 | 1,055 | 2,174 |
| **Gender (N=2,146)** |  |  |  |
| Female | 514 (46.4%) | 502 (48.4%) | 1,016 (47.3%) |
| Male | 567 (51.1%) | 515 (49.7%) | 1,082 (50.4%) |
| Other<sup>†</sup> | 28 (2.5%) | 20 (1.9%) | 48 (2.3%) |
| **Age (N=2,174)** | 14.5 (1.5) | 14.4 (1.5) | 14.4 (1.5) |
| **Planning to study beyond high school (N=2,045)** |  |  |  |
| Yes | 830 (79.2%) | 765 (76.7%) | 1,595 (78.0%) |
| No | 218 (20.8%) | 232 (23.3%) | 450 (22.0%) |
| **Missed school in the last month<sup>#</sup> (N=2,070)** |  |  |  |
| None | 704 (66.3%) | 709 (70.3%) | 1,413 (68.3%) |
| 1 or 2 days | 233 (21.9%) | 184 (18.2%) | 417 (20.1%) |
| > 2 days | 125 (11.8%) | 115 (11.4%) | 240 (11.6%) |
| **Physical violence by parents or figures of authority (N=1,918)** |  |  |  |
| Yes | 102 (10.1%) | 116 (12.8%) | 218 (11.4%) |
| No | 912 (89.9%) | 788 (87.2%) | 1,700 (88.6%) |
| **Forced sexual intercourse (N=1,953)** |  |  |  |
| Yes | 53 (5.1%) | 59 (6.4%) | 112 (5.7%) |
| No | 983 (94.9%) | 858 (93.6%) | 1,841 (94.3%) |
| **Ever had sexual intercourse for money or a gift (N=2,054)** |  |  |  |
| Yes | 49 (4.6%) | 65 (6.5%) | 114 (5.6%) |
| No | 1,011 (95.4%) | 929 (93.5%) | 1,940 (94.4%) |
| **Sexual intercourse for money or a gift in the last 6 months (N=2,048)** |  |  |  |
| Yes | 36 (3.4%) | 34 (3.4%) | 70 (3.4%) |
| No | 1,025 (96.6%) | 953 (96.6%) | 1,978 (96.6%) |
| **CKAQ-R score (N=1,779)** | 6.5 (2.1) | 6.6 (2.3) | 6.6 (2.2) |
| **Knowledge of risks of online sex score (N=1,616)** | 9.9 (3.2) | 10.0 (3.1) | 10.0 (3.1) |
| **Would seek help in cases of online abuse (N=2,010)** | 742 (70.7%) | 674 (70.1%) | 1,416 (70.4%) |
| **Participated in sexual abuse prevention activities (N=2,089)** | 277 (25.5%) | 283 (28.2%) | 560 (26.8%) |

Data are n (%) or mean (SD).
CKAQ-R=Children's Knowledge of Abuse Questionnaire – Revised; SD=Standard Deviation
\* Total number of students fitting eligibility criteria, as communicated by the school.
<sup>#</sup> Number of days reported by survey participants
<sup>†</sup> Others include transgender, non-binary, or those who respond 'other' when asked about their gender

Students’ average age was 14 years old, and 50.4% were male. More than one in twenty adolescents reported having experienced the most severe forms of sexual abuse and exploitation, with 5.7% reporting ever having sexual intercourse against their will, while 5.6% reported ever having sexual intercourse in exchange for money, gifts, or favors.

The mean CKAQ-R score was 6.6 at baseline (out of 12 questions), while the mean score for knowledge of risk associated with online sex was 10.0 (out of 15 questions). Around 26% of participants reported having participated in educational activities or classes on preventing or reacting to sexual abuse. All characteristics were well balanced between study arms.

One school from the control arm withdrew from the study after the baseline survey.

### Intervention implementation

The intervention was implemented between May and August 2024. Data about the implementation of the intervention are reported in Table 2.

**Table 2.** Intervention implementation (N= 30 schools in GWV arm)

|  | **N (%)**<br>**Mean (SD)** | **Endline**<br>**CKAQ-R** | **P-value<sup>§</sup>** |
| --- | --- | --- | --- |
| **School received GWV training** | 27 (90%) |  |  |
| **Number of teachers trained\*** | 1.7 (0.9) |  |  |
| **Number of GWV components implemented<sup>†</sup>** | 3.7 (2.7) |  |  |
| **School implementation level<sup>‡</sup>** |  |  |  |
| Not implemented or no data <sup>¶</sup> | 10 (33.3%) | 7.3 (0.4) | 0.559 |
| Lower (≤3 components) | 11 (30.0%) | 7.6 (0.7) |  |
| Higher (>3 components) | 9 (36.6%) | 6.9 (0.5) |  |

\* mean (SD) number of teachers recorded on training log. Based on 27 schools that attended training.
<sup>†</sup> mean (SD) number of GWV components used in class. Based on the teachers' survey, completed by 32 teachers in 20 schools.
<sup>‡</sup> Defined according to the number of GWV components used in class. Based on the teachers' survey, completed by 32 teachers in 20 schools.
<sup>§</sup> p-value comparing the mean CKAQ-R score between the three implementation levels. Adjusted for strata and baseline CKAQ-R.
<sup>¶</sup> Two schools (6.7%) did not receive the intervention, and eight (26.6%) received the intervention but did not complete the implementation survey

Among the 30 schools randomised to receive the GWV intervention, 3 (10%) schools enrolled teachers in GWV who did not attend the training. In the remaining 27 schools, an average of 1.7 teachers participated in the GWV training, with a range of 1 to 4 per school.

Implementation data were derived from a survey conducted with teachers in August 2024, following the GWV training sessions and prior to the endline assessment. The survey was completed by 32 teachers from 20 schools. On average, they reported using 3.7 GWV components in their classes. Four teachers (12%) reported not having implemented any GWV components. The reasons for this included not receiving the materials, difficulty in determining the best strategy to address such a sensitive subject, lack of time, and plans to implement the components in the future.

### Endline and outcomes

A small proportion (2.8%) of parents/guardians of eligible students refused their children’s participation in the survey. The endline survey was conducted in September 2024 and completed by 2,139 students from 59 schools (36.2 students per school on average). Participants’ characteristics are reported in Table 3.

**Table 3.** endline characteristics.

|  | Control | GWV | All |
| --- | --- | --- | --- |
| **School characteristics** |  |  |  |
| **Number of schools** | 29 | 30 | 59 |
| **Number of respondents per school** | 36.2 (9.7) | 36.3 (7.3) | 36.2 (8.5) |
| **Student characteristics** |  |  |  |
| **Number of student respondents** | 1,050 | 1,089 | 2,139 |
| **Gender (N=2,115)** |  |  |  |
| Female | 517 (49.9%) | 531 (49.2%) | 1,048 (49.6%) |
| Male | 501 (48.4%) | 529 (49.0%) | 1,030 (48.7%) |
| Other* | 18 (1.7%) | 19 (1.8%) | 37 (1.75%) |
| **Age (N=2,139)** | 14.7 (1.6) | 14.8 (1.5) | 14.8 (1.5) |

Data are n (%) or mean (SD).
\*Others include transgender, non-binary, or those who respond 'other' when asked about their gender.

The results for the primary and secondary outcomes are reported in Table 4.

**Table 4.** Primary and secondary outcomes.

|  | Control<br>(n=1,050) | GWV<br>(n=1,089) | Comparison* |  |  |
| --- | --- | --- | --- | --- | --- |
|  | N (%)<br>Mean (SD) | N (%)<br>Mean (SD) | Adjusted Mean<br>difference, RR,<br>or RD | 95% CI | p-value |
| **Primary Outcome** |  |  |  |  |  |
| CKAQ-R<sup>†</sup> | 7.21 (1.8) | 7.33 (1.9) | 0.02 | -0.19 to 0.23 | 0.87 |
| **Secondary Outcomes** |  |  |  |  |  |
| Knowledge of risks<br>associated with<br>online sex | 11.51 (2.1) | 11.50 (2.1) | -0.12 | -0.34 to 0.11 | 0.313 |
| Seeking help in cases<br>of online abuse | 858 (81.7%) | 870 (79.9%) | RD=-2.5%<br>RR=0.96 | -6.7% to 1.7%<br>0.91 to 1.02 | 0.235 |
| Participation in<br>sexual abuse<br>prevention activities | 285 (27.1%) | 289 (26.5%) | RD=-0.8%<br>RR=0.93 | -5.1% to 3.5%<br>0.78 to 1.11 | 0.717 |

CKAQ-R=Children's Knowledge of Abuse Questionnaire - Revised; CI=Confidence interval; RD= Risk difference; RR=Risk ratio \*All comparisons stratified by strata. For CKAQ-R and knowledge of online risks, analyses also adjusted for school mean baseline CKAQ-R and knowledge of online risks scores, respectively.

There was no missing data for the primary or secondary outcomes among the 2,139 students who completed the survey. The mean CKAQ-R score at endline was 7.33 in the intervention schools, compared to 7.21 in the control schools, with no statistically significant difference between arms (adjusted mean difference (AMD): 0.02, 95%CI −0.19 to 0.23, p=0.87).

There was no statistically significant difference between the two arms for the four secondary outcomes. The knowledge risk score for online sexual exploitation was 11.50 in GWV schools compared to 11.51 in control schools (adjusted mean difference [AMD]: −0.12, 95% CI −0.34 to 0.11, p=0.313). Additionally, 79.9% of students in GWV schools reported that they would seek help if their intimacy were exposed or if they were sexually harassed online, compared to 81.7% in control schools (risk difference [RD]: −2.5%, 95% CI: −6.7% to 1.7%, p=0.235). Furthermore, 26.5% of students in GWV schools reported having participated in educational activities or classes on preventing or responding to physical sexual abuse, compared to 27.1% in control schools (RD: −0.78%, 95% CI: −5.1% to 3.5%, p=0.717).

### Interaction

We examined the interaction between students’ gender and age regarding the intervention’s effect on the primary outcome. No evidence of interaction with age was found (see Table 5); however, there was borderline evidence of an interaction by gender, showing a greater difference between the arms in CKAQ-R scores for girls than for boys (interaction for boys vs girls: −0.34, 95% CI −0.65 to −0.03, p=0.032).

**Table 5.** interaction analysis by gender and age.

|  | Control | GWV | Mean difference* | Interaction* | Interaction p-value* |
| --- | --- | --- | --- | --- | --- |
| **By gender<sup>‡</sup>** |  |  |  |  |  |
| Female (n=1,053) | 7.29 | 7.56 | 0.18 |  |  |
| Male (n=1,034) | 7.14 | 7.08 | -0.15 | -0.34 (-0.65 to -0.03) | 0.032 |
| **By age** |  |  |  |  |  |
| <15 years (n=815) | 6.86 | 6.93 | 0.00 |  |  |
| ≥15 years (n=1,324) | 7.43 | 7.56 | 0.05 | 0.05 (-0.31 to 0.42) | 0.201<sup>†</sup> |

Data are mean CKAQ-R at endline
\*Comparing GWV arm to the control arm, using a mixed-model, adjusted for strata and school mean baseline CKAQ-R.
<sup>†</sup> Test for interaction assuming a linear interaction with age.
<sup>‡</sup> Transgender female or male includes female or male, respectively. Non-binary and 'other' are not included in this analysis.

### Dose-response

In a prespecified exploratory analysis, the 30 schools in the GWV arm were classified into three groups based on teachers’ responses to the implementation survey (Table 3). Ten schools (33.3%) either did not implement the intervention or lacked implementation data; 11 schools (30.0%) reported implementing an average of 3 or fewer GWV components; and 9 schools (36.6%) reported having implemented more than 3 components. Supplementary Material 2 details the components implemented by the teachers trained in the GWV methodology. There was no significant difference in adjusted mean CKAQ-R at endline between these 3 groups (p=0.559, Table 2)

## DISCUSSION

This trial found no evidence that the Growing Up Without Violence curriculum improved students’ knowledge of risks related to sexual abuse and exploitation. We also did not observe any effect on self-reported knowledge of online sexual risks, intentions to seek help in cases of online abuse, or participation in activities focused on sexual abuse prevention.

While it is difficult to pinpoint the exact reasons for the lack of intervention effects observed in this trial, several factors could contribute to this outcome. These findings may reflect flaws in the GWV’s rationale and assumptions (e.g., the belief that changes in knowledge would lead to changes in behaviour), ineffective implementation strategies, or contextual challenges to implementation. Furthermore, the trial design may not have adequately captured certain effects, such as longer-term impacts or effects on other outcomes.

Previous studies of school-based prevention of child sexual abuse demonstrated an overall effectiveness of programmes targeting children’s knowledge of risks and protective behaviours (11, 33). Although the findings from these studies are not generalisable to the GWV intervention design or our study context, they indicate that school-based interventions can affect students’ CSAE knowledge levels. Previous findings from a non-published GWV qualitative evaluation in a different Brazilian municipality showed that teachers considered school-based actions on CSAE urgent and valued the GWV content. However, they faced many barriers to implementation, including limited resources and competing priorities for teachers’ time.

Similarly, we hypothesize that the null results from our trial may be partially due to implementation challenges and contextual barriers affecting effectiveness. These include a shortened implementation timeline, low school engagement, high student absenteeism, and obstacles to positive teacher-student relationships. We discuss these factors in more detail below.

### Reduced timeline for implementation

Extended negotiations with municipal and state authorities to finalize agreements with Canal Futura and secure authorization for school-based research activities reduced the implementation timeline from 8 months to just 3 months. This shortened interval between baseline and endline limited opportunities for effective GWV implementation, resulting in a low dose of the intervention being delivered. Additionally, students who received the curriculum had their chances for reflection, abstract conceptualization, and active experimentation with GWV concepts significantly constrained.

Despite this reduced timeline, our findings show that most teachers (66.6%) reported delivering at least some of the GWV components. Conversely, only 26.5% of students recalled ever participating in CSAE prevention activities. This discrepancy may suggest that, particularly in larger schools, the number of students who have direct contact with trained teachers is limited. Implementing strategies to ensure coverage in all classrooms could help expand the intervention’s reach.

### Low school engagement

The reduced timeline of the intervention may have provided insufficient time for teachers to conduct GWV activities alongside the demanding requirements of the Brazilian national curriculum. Low engagement could also stem from cultural and religious barriers to the intervention’s content. For instance, one school withdrew from the intervention, deeming the survey questions on sex and violence inappropriate for children and citing parental opposition. Resistance to GWV might also relate to teachers feeling uncomfortable with the content or holding attitudes that contradict the intervention’s objectives. However, lack of engagement alone cannot fully account for the trial’s findings, as we did not observe improved outcomes in schools with higher implementation levels, nor did we see an increase in students recalling exposure to CSAE activities in intervention schools compared to control schools.

### Students’ high absenteeism

High levels of absenteeism may have adversely affected students’ engagement with the intervention, resulting in a low dose of exposure. During both baseline and endline fieldwork, our teams found that nearly a third of the sampled students were absent from school. Additionally, baseline findings indicated that one in five students reported interrupting their studies for more than a month for reasons unrelated to the COVID-19 pandemic. This high absenteeism reduces children’s likelihood of exposure to GWV activities, which, in turn, affects their recall of intervention activities and content learning. Moreover, research indicates that school absenteeism is linked to increased youth vulnerability, including higher rates of potentially harmful sexual behaviours, psychiatric disorders, externalizing behaviours, youth crime, and alcohol and substance misuse (34). Several of these risks associated with absenteeism overlap with risks of sexual exploitation, suggesting that some of the students who would benefit most from interventions like GWV are also more likely to miss school.

### Barriers impeding positive teacher-student relationships

Pedagogical research indicates that a positive educational environment is central to effective learning, critical thinking, and students’ development (35). Findings from our baseline survey revealed that a high proportion of students felt they were they were not often allowed to express their opinions (46.0%) or ask questions in the classroom (43.0%). Additionally, more than two-fifths of surveyed students believed their peers frequently disrupted classes, which may have hindered teachers’ ability to deliver content effectively.

While most students perceived their teachers as motivated (80.5%), the majority reported that teachers rarely engaged with them (70.6%) and showed no interest in their lives outside school (57.5%). These findings indicate a lack of equitable dialogue and engagement between some students and their teachers, suggesting that students may not feel they have a voice in their relationships with their educators. This could undermine trust and engagement in CSAE prevention activities addressing particularly sensitive topics.

In interventions aimed at promoting normative change, the absence of space for dialogue and open discussion may serve as a significant barrier (36). Furthermore, if students’ perceptions are accurate and some teachers demonstrate a lack of interest in students’ out-of-school experiences, these teachers may be less likely to engage meaningfully with the intervention topics or proactively identify students in need of support. Lastly, students’ disruptive behaviour could further impede teachers’ ability to deliver GWV content effectively.

### Strengths and Limitations

To our knowledge, ours was the first rigorous large-scale evaluation of a school-based intervention to prevent CSAE in Brazil. The analysis plan was pre-specified, with a limited number of comparisons. Furthermore, all the data and analysis had been prepared before unmasking, limiting the risk of chance finding.

The study, however, had several limitations, outlined below.

#### Generalisability

We conducted the study in two municipalities in Pernambuco State, in the Northeast of Brazil. Pernambuco State is where the research sponsor, the Freedom Fund, has a hotspot for CSAE intervention. Canal Futura selected Cabo and Jaboatão as implementation sites because they had good connections with the local authorities and had not implemented GWV there recently. Although we have no reason to believe the trial results would differ in other regions of the country, we cannot be sure they are generalisable.

#### Challenges to implement GWV as planned

Delays in obtaining authorization from local authorities significantly shortened the timeframe between baseline and endline assessments. Consequently, the trial was limited to a restricted implementation period for GWV activities, capturing only 3 of the originally planned 8 months. Given the sensitive nature of the topic and the deeply ingrained social norms that support or excuse CSAE in the research context, it is likely that the GWV implementation period was insufficient to yield meaningful results. Additionally, there is insufficient data to evaluate the quality or consistency of implementation, particularly given the intervention’s unstructured nature. We conducted an initial online survey with all teachers enrolled in the intervention, but low participation (40%) reduced the utility of these data.

#### Statistical power

Although the number of respondents was lower than anticipated due to absenteeism, the study enrolled a substantial number of schools and maintained sufficient power to detect a moderate to large effect size. With an intraclass correlation coefficient (ICC) of 0.06 and an average cluster size of 36 students per school, the power to detect an effect size of 0.25 remained above 90%. The confidence interval for the primary outcome dates any significant effect, although we cannot entirely rule out a small benefit.

## CONCLUSION

This study did not find an impact of GWV on the intended outcomes. Several factors may contribute to this result: the intervention might have been implemented with insufficient fidelity or intensity to produce measurable change, the true effect of GWV could be small and fall below the study’s detectable threshold, or the intervention may simply lack effectiveness. Based on our findings, we cannot draw definitive conclusions about the intervention’s potential to effect change.

Future implementation of the GWV and similar initiatives should prioritise strategies to enhance and monitor the quality and intensity of delivery in both intervention and control groups. For example, future impact evaluations should ensure adequate investment in comprehensive process evaluations. Additionally, implementation could be strengthened by training multiple educators per school or employing training-of-trainers approaches to reinforce the content and broaden outreach.

CSAE is a pervasive issue that has devastating effects on children’s health, well-being, and development. Previous evaluations of school-based CSAE prevention programs have yielded mixed results. To ensure that resources are allocated to effective strategies, further rigorous evaluations and implementation studies are essential.

## Data Availability

All files will be publicly available from a public data repository.

## ACKNOWLEDGEMENTS

We would like to acknowledge the unwavering support of Cinthia Sarinho and Priscila Pereira from Canal Futura during the planning and implementation of the evaluation, as well as in the interpretation of findings. Their invaluable contextual knowledge and professional expertise have been instrumental in ensuring the quality and relevance of our study.

Funding was made possible by the United States Department of State under the terms of Cooperative Agreement No. SSJTIP20CA0026, through the Human Trafficking Research Initiative, managed by Innovations for Poverty Action.

This research was funded by a grant from the United States Department of State. The opinions, findings, and conclusions herein are those of the authors and do not necessarily reflect those of the United States Department of State.

## AUTHORS CONTRIBUTIONS

LKi was responsible for the conceptualisation of the research, methodological design, and overall supervision. LKi, YL, and EA handled funding acquisition. LKi wrote the first draft of the manuscript and integrated the co-authors’ feedback. BL led methodological development, designed the formal analysis, supervised the statistical analysis, and contributed to the first draft of the manuscript. ML conducted the formal statistical analysis, managed related data curation, and reviewed and contributed to the final version of the manuscript. AP and HP supervised all research activities in Brazil and contributed to the project’s conceptualisation and administration. Both reviewed the manuscript and provided important contributions. AI and MP contributed to project conceptualisation, data curation, resource development, administration, and data analysis. Both reviewed the manuscript and offered meaningful contributions. HS, YL, and EA had critical roles in the project conceptualisation and provided significant input to the manuscript. HS, YL, and EA played critical roles in project conceptualisation and provided significant input to the manuscript. LKe and CK supervised ML’s role and offered critical contributions to the paper. DB and MB had key roles in project administration and resource provision and contributed meaningfully to the manuscript.

## CONFLICT OF INTEREST

The authors of this manuscript have no conflicts of interest to declare.

## Notes

### Competing Interest Statement

The authors have declared no competing interest.

### Clinical Trial

AEARCTR-0011299, https://www.socialscienceregistry.org/trials/11299

### Clinical Protocols

https://pubmed.ncbi.nlm.nih.gov/39800276/

https://figshare.com/articles/preprint/Growing_Up_Without_Violence_Trial_-_Statistical_Analysis_Plan/28024862?file=51171968

### Author Declarations

Ethical approval for this research was granted by the Research Ethics Committee at the University College London (Project ID/Title: 24085/001), and the Brazilian National Research Ethics Committee (CAAE: 66827122.0.0000.5206).

